# Spatiotemporal Characteristics of Cerebrospinal Fluid Flow Across the Craniospinal Axis Using Cine Phase-Contrast MRI: A Ventral-Dorsal Dual-Region Flow Pattern in the Spinal Subarachnoid Space

**DOI:** 10.64898/2026.07.29.26359135

**Authors:** Liying Sun, Le He, Zhimo Jian, Tiange Lu, Suhua Miao, Rongsong Zhou, Tiemin Li, Mingyuan Yan, Yuqi Zhang, Yajun Yin, Yu Ma

**Author notes:** These authors contributed equally to this work (Liying Sun, Le He). **Correspondence:** Yu Ma, Department of Neurosurgery, Tsinghua University Yuquan Hospital (Tsinghua University Hospital of Integrated Traditional Chinese and Western Medicine), Beijing, China.; Yajun Yin, Tsinghua University, Beijing, China.; Yuqi Zhang, Department of Neurosurgery, Tsinghua University Yuquan Hospital (Tsinghua University Hospital of Integrated Traditional Chinese and Western Medicine), Beijing, China.

## Abstract

**Background:** Cerebrospinal fluid (CSF) circulation is important for maintaining homeostasis of the central nervous system. Previous studies have largely focused on the ventricular system, the craniocervical junction, or local spinal segments, leaving the overall and spatially heterogeneous characteristics of CSF flow across the craniospinal axis insufficiently characterized. The spinal subarachnoid space (SAS) is often treated as a homogeneous annular compartment surrounding the spinal cord, an approach that may obscure directional differences among its internal regions.

**Methods:** This single-center, exploratory, prospective imaging study enrolled 15 healthy volunteers. All participants underwent 3.0-T electrocardiography-gated two-dimensional cine phase-contrast magnetic resonance imaging (Cine PC-MRI) and high-resolution T2-weighted imaging. CSF was evaluated at the level of the cerebral aqueduct outlet/fourth-ventricle inlet, C1-C2, C5-C6, T5-T6, L1-L2, and the lumbar cistern. Region-of-interest (ROI)-based quantitative analysis using Q-Flow software recorded mean velocity, absolute peak velocity, and directional peak velocity.

**Results:** Multiplanar Cine PC-MRI showed that CSF phase signals within the spinal SAS were not uniformly distributed but formed two principal flow regions, ventral and dorsal. Mean velocity and absolute peak velocity were similar between the ventral and dorsal regions, whereas directional peak velocity differed (1.50 ± 2.98 cm/s vs. -0.38 ± 3.23 cm/s, P = 0.036). High-resolution T2-weighted imaging showed denticulate ligaments, nerve roots, and associated fibrous connective tissue in the lateral transition zones between the two regions.

**Conclusions:** In healthy adults, CSF flow in the spinal SAS was not synchronous motion within a single homogeneous compartment; rather, it showed longitudinal oscillatory flow in ventral and dorsal regions coupled to the cardiac cycle. These findings provide preliminary in vivo evidence for studies of CSF hydrodynamics across the craniospinal axis and an imaging basis for investigating CSF circulation disturbances in conditions such as hydrocephalus, Chiari malformation, syringomyelia, and arachnoid adhesions.

## Background

This study aimed to observe CSF flow characteristics across the craniospinal axis. CSF circulation contributes to intracranial pressure regulation, maintenance of the perineural microenvironment, clearance of metabolic products, and solute transport [1, 2]. CSF circulation disturbances have been associated with hydrocephalus, Chiari malformation, syringomyelia, and neurodegenerative diseases [3]. Accordingly, investigation of CSF circulation has anatomical relevance and is also pertinent to studies of disease mechanisms and treatment evaluation in disorders of the central nervous system.

The conventional model of CSF circulation focuses on the pathway of choroid plexus secretion, flow through the ventricular system, and absorption by arachnoid granulations [4, 5]. This model provides an important basis for understanding the ventricular system, intracranial pressure regulation, and disorders such as hydrocephalus. However, it focuses primarily on intracranial CSF circulation and provides limited information about the directionality, periodicity, and regional distribution of CSF flow within the spinal SAS. The spinal SAS is not simply an extension of intracranial CSF circulation; it is a continuous space connecting the intracranial SAS, perispinal spaces, and the lumbar cistern. An understanding of CSF hydrodynamics based only on the ventricular system or a single craniocervical level is unlikely to reflect the overall characteristics of craniospinal CSF flow [3], and there is no consensus on the spatial organization of spinal CSF flow. Previous studies have several limitations: (1) PC-MRI observations have commonly been made at the cerebral aqueduct, craniocervical junction, or a single spinal segment; (2) the spinal SAS has commonly been assumed to be a single annular compartment surrounding the spinal cord; and (3) the entire spinal SAS has been delineated as one ROI to obtain global parameters. However, a global ROI is a measurement and modeling simplification. Previous work suggests that spinal CSF flow is pulsatile, bidirectional, and spatially heterogeneous [6, 7]. Hydrodynamic averages derived from a global ROI may cancel signals with opposing directions, attenuate regional signals, and thereby limit investigation of spatial heterogeneity [8].

To address these issues, we used electrocardiography-gated Cine PC-MRI with Q-Flow quantitative analysis at six standardized acquisition planes: the cerebral aqueduct outlet/fourth-ventricle inlet, C1-C2, C5-C6, T5-T6, L1-L2, and the lumbar cistern. This enabled multiplanar, dynamic, and quantitative observation of CSF flow across the craniospinal axis. Rather than applying a single homogeneous-compartment assumption to the spinal SAS, ROIs were separately delineated for the reproducibly identifiable ventral and dorsal principal signal regions on phase images, while retaining directional information. We aimed to describe the spatial distribution of CSF flow across multiple planes, assess whether ventral and dorsal dual regions can be identified consistently within the spinal SAS, and compare their flow characteristics.

## Methods

### Study design and participants

This prospective study enrolled 15 healthy volunteers without a history of neurological disease or contraindications to MRI. Healthy participants were selected to minimize the influence of disease-related factors on CSF hydrodynamics and to obtain baseline observational data under healthy conditions. All participants fasted and abstained from water for at least 4 h before scanning to reduce the effect of gastrointestinal motion on lumbosacral imaging. The study was approved by the Medical Ethics Committee of Tsinghua University Yuquan Hospital (approval no. 2024KY014), and all participants provided written informed consent.

#### MRI acquisition and standardized multiplanar Cine PC-MRI strategy

The methodological innovation of this study was not the development of a new MRI pulse sequence. Instead, in view of the low-velocity and pulsatile characteristics of CSF flow, we extended conventional two-dimensional cine phase-contrast magnetic resonance imaging (Cine PC-MRI) from the commonly used single local-plane measurement to a standardized multiplanar sampling framework covering the craniospinal axis. This design used fixed anatomical landmarks, a uniform definition of velocity direction, a uniform velocity encoding value, and a uniform post-processing workflow, allowing CSF velocity parameters to be compared across segments and regions within the same axial plane.

All MRI examinations were performed on a 3.0-T whole-body scanner (Philips Ingenia CX, Best, the Netherlands) with a 16-channel body coil. Participants were positioned supine, head first. Electrocardiography-gated two-dimensional Cine PC-MRI was acquired at six anatomically defined levels that were consistent across participants: the cerebral aqueduct outlet/fourth-ventricle inlet, C1-C2, C5-C6, T5-T6, L1-L2, and the lumbar cistern. Imaging parameters were as follows: field of view, 150 mm x 150 mm; matrix, 256 x 256; nominal in-plane spatial resolution, 0.59 mm x 0.59 mm; slice thickness, 8 mm; repetition time/echo time (TR/TE), 13/8.5 ms; 15 phases per cardiac cycle; and velocity encoding (VENC), 6 cm/s. Cranial flow was defined as positive and caudal flow as negative.

#### Multiplanar anatomical localization strategy

Previous CSF hydrodynamic studies have often selected a single observation plane, such as the cerebral aqueduct or craniocervical junction. A single plane can provide local information but is insufficient to reflect flow characteristics across the whole spinal cord [3, 7]. We selected six standardized acquisition planes along the craniospinal axis to observe CSF motion continuously from the intracranial compartment to the lumbar cistern and to compare flow distributions across spinal segments.

**Table 1.**
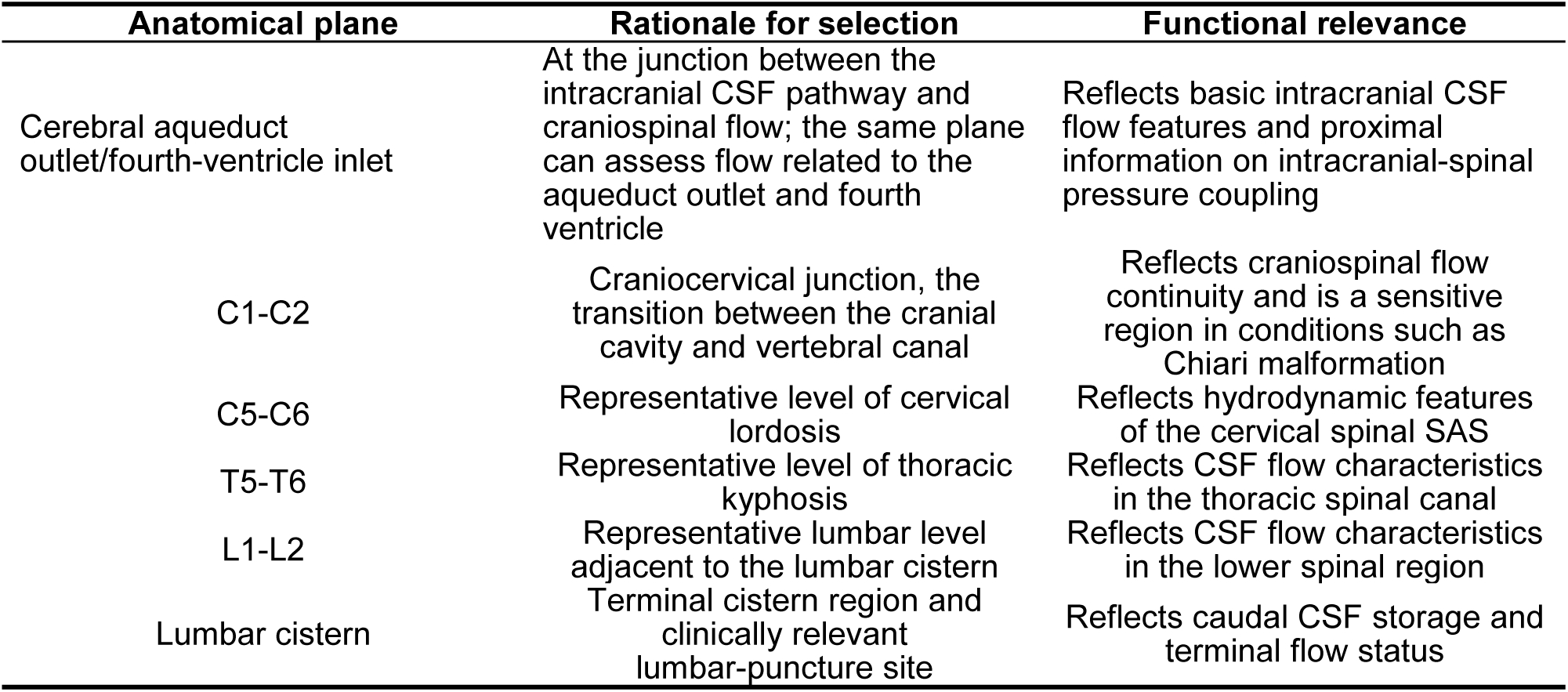
Multiplanar anatomical localization strategy.

### Image post-processing, ROI definition, and statistical analysis

The focus of image post-processing was to preserve regional flow information in the spinal SAS that might be averaged by a global ROI. ROIs were delineated by integrating axial anatomy with the distribution of velocity signals on phase-contrast images to compare directional flow characteristics between ventral and dorsal regions.

All imaging data were analyzed on a Philips IntelliSpace Portal post-processing workstation. Q-Flow software was used for ROI delineation and parameter extraction at the prespecified anatomical levels. On axial spinal phase images, CSF velocity-encoded signals were mainly distributed in ventral and dorsal regions, with relatively stable ancillary flow-signal regions visible laterally. ROI boundaries were determined according to the spatial distribution of flow signals, anatomical continuity, and the morphology of the SAS around the spinal cord. The lateral ancillary signal regions assisted in defining the boundary of the corresponding ventral or dorsal ROI and were not analyzed as independent regions. The spinal SAS was divided into two principal ROIs, ventral and dorsal. Mean velocity, absolute peak velocity, and directional peak velocity were extracted for each ROI. Directional peak velocity retained the positive or negative sign of velocity encoding to reflect flow direction, whereas absolute peak velocity described velocity magnitude without regard to direction.

Two physicians with experience in central nervous system imaging analysis independently performed ROI delineation and parameter measurements. Interobserver agreement was assessed using the intraclass correlation coefficient (ICC).

Statistical analyses were performed with IBM SPSS Statistics 27.0. Comparisons between ventral and dorsal ROIs used the participant as the unit of analysis. Measurements from the corresponding spinal segments were first summarized for each participant to derive participant-level representative values for the ventral and dorsal regions; paired comparisons were then performed to reduce the risk of pseudoreplication from treating multiple planes within the same participant as independent observations. Continuous variables are presented as mean ± standard deviation or median (interquartile range), according to their distribution. Paired data from ventral and dorsal regions were compared using a paired t test or Wilcoxon signed-rank test, according to distributional characteristics. All tests were two-sided, and P < 0.05 was considered statistically significant.

## Results

### Participant characteristics and measurement agreement

Fifteen healthy volunteers were included. The mean age was 39.53 ± 14.09 years; five participants were male and ten were female. Median height was 163 cm (interquartile range, 159-172 cm), and mean weight was 60.9 ± 11.52 kg. Overall agreement between the Q-Flow parameters measured by the two observers was good (ICC > 0.75).

**Table 2.** Baseline characteristics of study participants (n = 15)

| Characteristic | Value |
| --- | --- |
| Age, years | 39.53±14.09 |
| Sex, male/female | 5/10 (33.3%/66.7%) |
| Height, cm | 163 (159, 172) |
| Weight, kg | 60.9±11.52 |

### Multiplanar imaging shows spatial heterogeneity of spinal CSF flow

Cine PC-MRI was used to observe CSF flow at standardized planes from the cerebral aqueduct outlet/fourth-ventricle inlet to the lumbar cistern (Figure 3). Case-by-case review of all participants and planes showed a nonuniform axial distribution of CSF velocity-encoded phase signals within the SAS at the four spinal segmental levels of C1-C2, C5-C6, T5-T6, and L1-L2. Signals were mainly located ventral and dorsal to the spinal cord. Phase-signal intensity and direction differed between the two regions and showed an overall consistent ventral-dorsal partitioning trend across participants and spinal segments, suggesting relatively stable spatial heterogeneity of CSF flow (Figure 5). No consistent ventral-dorsal regional separation was observed in the lumbar cistern (Figure 5F). Anatomically, this region contains freely distributed cauda equina nerve roots rather than a spinal cord with a clearly defined axial contour. Compared with the spinal segments, the SAS in the lumbar cistern is relatively wide, nerve roots are more dispersed, and there is no stable ventral-dorsal spatial partition based on the spinal cord and its surrounding supporting structures. Accordingly, the lumbar cistern did not show a ventral-dorsal partition similar to that of the spinal segments. Small lateral ancillary signal regions were visible near nerve-root-related structures in some participants. The occurrence, extent, and phase characteristics of these signals varied between individuals; their anatomical basis and physiological significance cannot be determined directly from this study and may be compared in larger samples incorporating healthy controls and disease cohorts.

**Figure 1.**
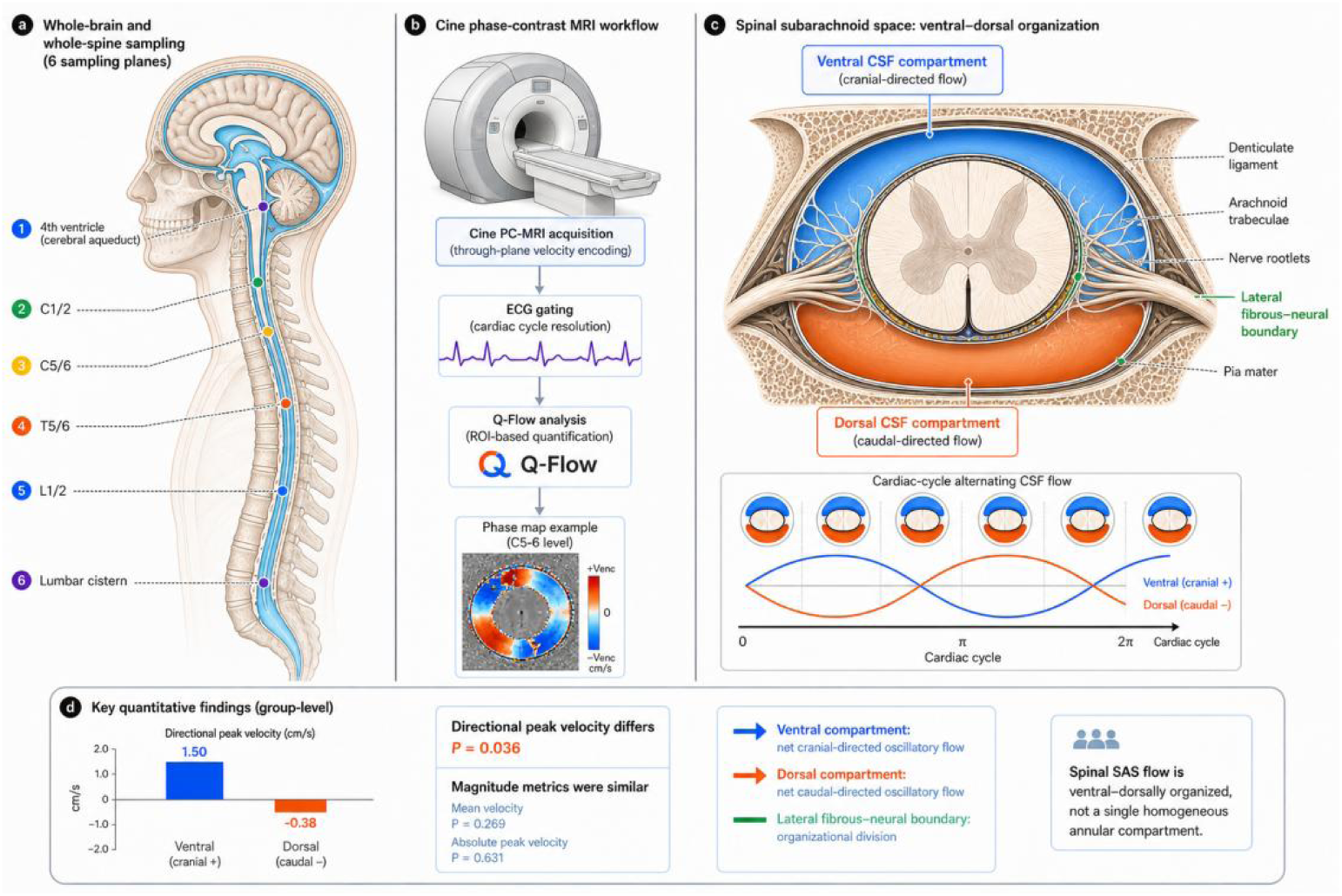
Observation workflow for CSF flow across the craniospinal axis and schematic of the ventral-dorsal dual-region flow pattern in the spinal SAS. (a) Standardized multiplanar Cine phase-contrast MRI (Cine PC-MRI) sampling strategy. Six anatomically defined levels were measured along the craniospinal axis: the cerebral aqueduct outlet/fourth-ventricle inlet, C1-C2, C5-C6, T5-T6, L1-L2, and the lumbar cistern. (b) Cine PC-MRI acquisition and quantitative analysis workflow. Through-plane velocity encoding and electrocardiography gating were used to resolve CSF motion across 15 phases of one cardiac cycle; ROI-based quantitative analysis was then performed with Q-Flow. The example shows a phase-contrast velocity map at C5-C6; cranial flow was defined as positive and caudal flow as negative. (c) Conceptual model of regional CSF flow in the ventral and dorsal spinal SAS. Cranially directed velocity signals predominate ventrally, whereas caudally directed signals predominate dorsally. Denticulate ligaments, arachnoid trabeculae, nerve rootlets, pia mater, and lateral fibrous-neural boundaries may form transition zones for this regional spatial separation. The curves below schematically depict cardiac-phase-related directional velocity changes. (d) Group-level quantitative results in healthy participants. Directional peak velocity differed between ventral and dorsal regions (1.50 cm/s vs. -0.38 cm/s, P = 0.036), whereas mean velocity (P = 0.269) and absolute peak velocity (P = 0.631) did not differ significantly. These findings support a regional ventral-dorsal CSF flow phenotype in the spinal SAS rather than a single, homogeneous annular flow space.

**Figure 2.**
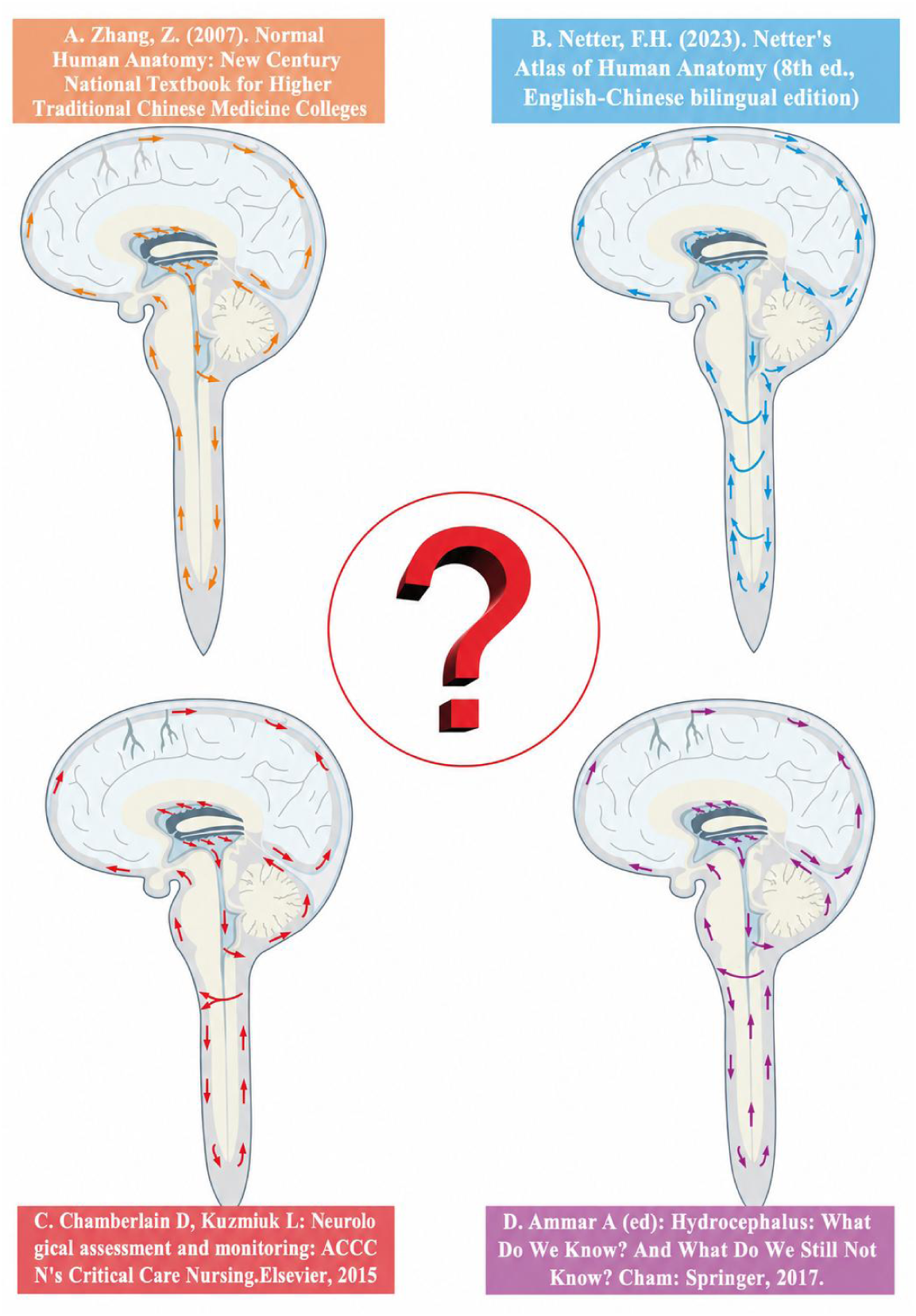
Schematic of previous CSF circulation models. Previous diagrams have described intracranial CSF circulation pathways relatively consistently, whereas the direction and spatial distribution of CSF flow in the spinal region have been described inconsistently. This highlights the need to reassess CSF hydrodynamic characteristics from a craniospinal perspective.

**Figure 3.**
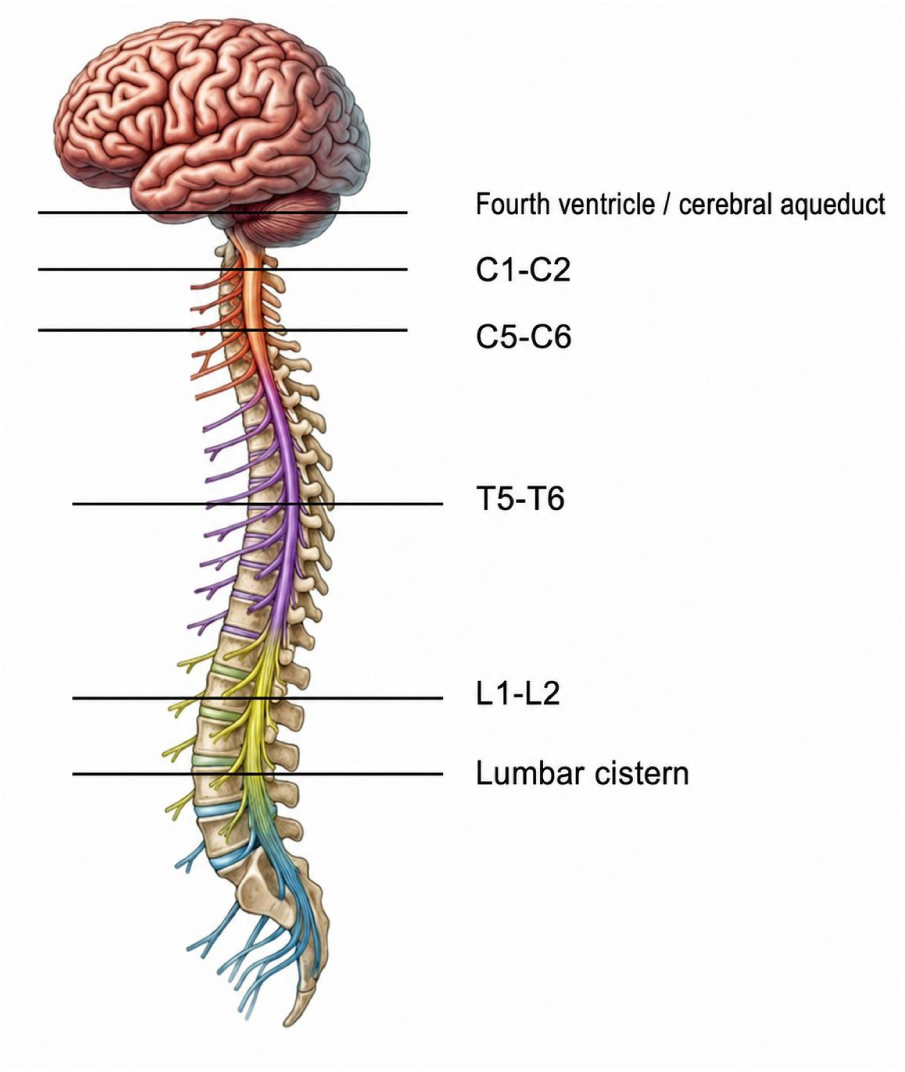
Six standardized acquisition planes for CSF hydrodynamic assessment. The planes were the cerebral aqueduct outlet/fourth-ventricle inlet, C1-C2, C5-C6, T5-T6, L1-L2, and the lumbar cistern, allowing continuous assessment of CSF flow from the intracranial compartment to the caudal spinal region.

**Figure 4.**
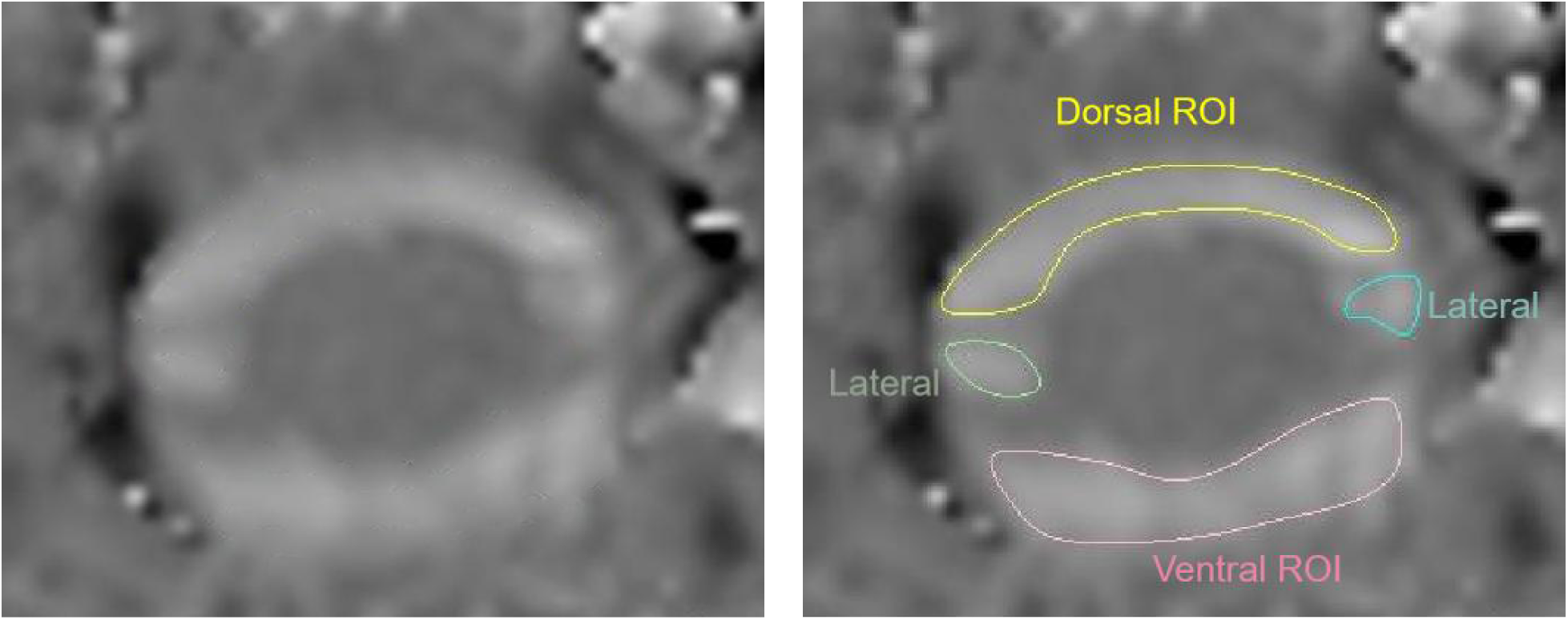
Schematic ROI delineation of ventral, dorsal, and lateral ancillary regions in the spinal SAS.

**Figure 5.**
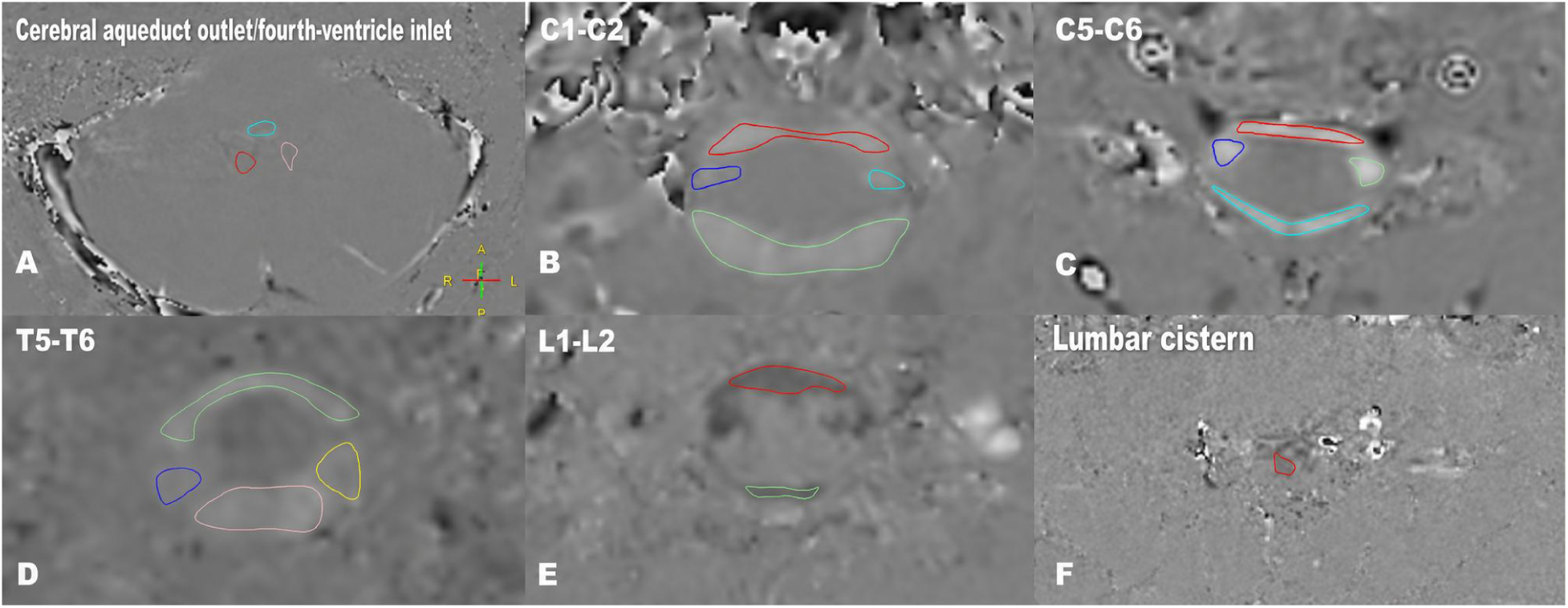
Representative Cine PC-MRI velocity-encoded phase images at six standardized acquisition planes. (A) Cerebral aqueduct outlet/fourth-ventricle inlet; (B) C1-C2; (C) C5-C6; (D) T5-T6; (E) L1-L2; and (F) lumbar cistern. At the four spinal segmental levels (B-E), CSF velocity-encoded phase signals were mainly distributed ventral and dorsal to the spinal cord, with small lateral ancillary signal regions. The lumbar cistern (F) did not show a consistent ventral-dorsal phase-signal partition. Colored outlines illustrate the locations of the ventral, dorsal, and lateral ancillary ROIs.

### Ventral and dorsal flow magnitudes are similar but directional characteristics differ

Participant-level paired comparisons showed no statistically significant difference in mean velocity between the ventral and dorsal principal flow regions (0.09 ± 0.11 cm/s vs. 0.11 ± 0.12 cm/s, P = 0.269). Absolute peak velocity was also similar between the two regions (3.09 ± 1.15 cm/s vs. 3.00 ± 1.13 cm/s, P = 0.631). However, the directionality of peak velocity differed between the regions (1.50 ± 2.98 cm/s vs. -0.38 ± 3.23 cm/s, P = 0.036). These results indicate comparable flow magnitude but different directional behavior over the cardiac cycle.

**Table 3.** Comparison of CSF hydrodynamic parameters in ventral and dorsal spinal SAS regions.

| Measure | Ventral | Dorsal | Test statistic | P value |
| --- | --- | --- | --- | --- |
| Mean velocity, cm/s | 0.09±0.11 | 0.11±0.12 | W=143.0 | 0.269 |
| Absolute peak velocity, cm/s | 3.09±1.15 | 3.00±1.13 | t=0.486 | 0.631 |
| Directional peak velocity, cm/s | 1.50±2.98 | -0.38 ± 3.23 | t=2.195 | 0.036* |

### Sequential cardiac-phase images support antiphase ventral-dorsal flow

Representative multiphase PC-MRI images showed that phase signals in the ventral and dorsal spinal SAS regions changed over the cardiac cycle. These changes were synchronized with electrocardiography gating and were consistent with the oscillatory nature of CSF flow. The findings suggest longitudinal oscillations with different phase or direction in the ventral and dorsal regions during the cardiac cycle, differing from the conventional concept of single-compartment flow.

## Discussion

### Main findings: a craniospinal imaging framework extending the conventional CSF circulation model

CSF circulation has long been regarded as an important component of fluid homeostasis in the central nervous system. The conventional model proposes that CSF is produced mainly by the choroid plexus in the ventricular system, passes through the interventricular foramina, third ventricle, cerebral aqueduct, fourth ventricle and its outlets into the SAS, then flows along the brain and spinal cord surfaces and is reabsorbed into the venous system via arachnoid granulations [4, 5]. This model provides a basic framework for understanding hydrocephalus, intracranial pressure regulation, and ventricular disorders, and has made the cerebral aqueduct, fourth ventricle, and craniocervical junction common observation sites in PC-MRI studies [6, 14].

However, limitations of the conventional model have become increasingly apparent with the development of neuroimaging and CSF hydrodynamic research. CSF motion is not simply unidirectional; both cardiac and respiratory cycles can drive reciprocating CSF movement [15]. Perivascular spaces, meningeal lymphatic vessels, perineural pathways, and local tissue interfaces may also contribute to fluid exchange and solute transport [16-18]. Previous studies have often focused on the intracranial ventricular system, the cerebral aqueduct, or a single cervical spinal level, with limited systematic investigation of the continuity, directionality, and regional variation of CSF flow across the whole spinal cord [3, 12].

To address these issues, we incorporated the cerebral aqueduct outlet/fourth-ventricle inlet, C1-C2, C5-C6, T5-T6, L1-L2, and the lumbar cistern into a single standardized observation framework, creating a multiplanar method for observing CSF hydrodynamics from the intracranial compartment to the lumbar cistern. Unlike the conventional approach of analyzing the spinal SAS as a single annular compartment, this study retained regional information within the axial plane. Regional analysis showed a nonuniform distribution of CSF phase signals in the spinal SAS, mainly concentrated in ventral and dorsal regions. Mean velocity and absolute peak velocity were similar, suggesting broadly comparable flow magnitude, whereas directional peak velocity differed. Sequential cardiac-phase images further showed opposite trends in phase signals in the ventral and dorsal regions over the cardiac cycle. These results indicate that regional differences in CSF flow are expressed primarily in directional characteristics rather than in flow magnitude.

This study established an in vivo, multiplanar method for observing CSF hydrodynamics across the craniospinal axis from the intracranial compartment to the lumbar cistern. The findings suggest that the spinal SAS should not be viewed simply as a homogeneous annular flow channel [3, 11]. Within the axial plane, ventral and dorsal regions displayed different directional flow characteristics, suggesting regional spatial partitioning. These findings also suggest that the conventional practice of treating the spinal SAS as a single annular compartment may affect interpretation of spinal CSF flow, offering one possible explanation for inconsistency among previous research findings and unsatisfactory clinical outcomes.

### Novel application of conventional two-dimensional Cine PC-MRI

The innovation of this study lies not in developing a new MRI sequence but in extending the application of conventional two-dimensional Cine PC-MRI. Previous studies have commonly used PC-MRI to measure CSF hydrodynamic indices at the cerebral aqueduct, craniocervical junction, or a single spinal segment, mainly assessing CSF flow velocity and direction within one plane. We used uniform sampling at six standardized anatomical levels from the cerebral aqueduct outlet/fourth-ventricle inlet to the lumbar cistern and separately analyzed flow characteristics within ventral and dorsal ROIs on axial spinal images. This strategy extends local-plane measurements into a multiplanar study across the craniospinal axis and adds regional information within the axial plane. It may help identify directional differences that are averaged in global ROI analysis. Mean velocity and absolute peak velocity were similar in the ventral and dorsal regions, whereas directional peak velocity differed statistically (1.50 ± 2.98 cm/s and -0.38 ± 3.23 cm/s, respectively; P = 0.036).

Two-dimensional Cine PC-MRI was appropriate as the primary measurement technique for the flow characteristics and objective of this study. CSF in the SAS mainly exhibits low-velocity, cardiac-cycle-related pulsatile motion. Two-dimensional Cine PC-MRI directly encodes velocity through the imaging plane and is suitable for observing such low-velocity pulsatile signals. We set VENC to 6 cm/s to balance phase sensitivity to low CSF velocities against the risk of velocity aliasing. Combined with electrocardiography gating and Q-Flow quantitative analysis, this method provides directional velocity information at multiple phases of a cardiac cycle. Uniform plane localization, velocity-direction definition, VENC setting, and post-processing made measurements at different anatomical levels comparable. The method can compare CSF flow features across segments and differences between ventral and dorsal regions within the same axial spinal plane. In addition, Cine PC-MRI does not require contrast material and is noninvasive, with relatively straightforward acquisition and post-processing. Q-Flow provides standardized ROI-based quantitative results for between-group, between-segment, and between-region comparisons [6, 13].

By comparison, 4D Flow MRI can provide three-dimensional velocity vector fields over the cardiac cycle and permit retrospective reconstruction of arbitrary planes. It is suitable for observing complex directional motion, streamline patterns, and spatial relationships across planes in a continuous volume, and can also estimate derived parameters such as pressure gradients [9, 10]. However, when coverage is expanded to a large craniospinal region, 4D Flow MRI cannot easily optimize coverage, spatial resolution, temporal resolution, and signal-to-noise ratio simultaneously. Increasing the scan range generally prolongs acquisition and increases post-processing complexity. In a direct comparison of two-dimensional PC-MRI and 4D Flow MRI in a patient-specific in vitro Chiari I malformation model, one 4D Flow MRI acquisition required approximately 15 min, whereas two-dimensional PC-MRI required approximately 30 s per axial plane. After peak systolic CSF velocities were compared across nine axial planes, the overall standard deviation of 4D Flow measurements was higher than that of two-dimensional PC-MRI (1.83 vs. 1.04 cm/s) [10]. For low-velocity CSF signals, reduced signal-to-noise ratio, background phase offsets, respiratory motion, and body-motion artifacts may also affect quantitative results [11, 12].

The core objective of this study was to compare low-velocity through-plane CSF flow at multiple standardized anatomical levels and to identify directional differences between ventral and dorsal regions on axial spinal images. For this objective, two-dimensional Cine PC-MRI was well suited. Although it cannot provide three-dimensional velocity information throughout an entire volume, it can generate focused, repeatable, and comparable quantitative measurements at multiple fixed planes, making it suitable for analyzing regional and segmental features of low-velocity, pulsatile CSF flow. Thus, Cine PC-MRI can provide repeatable, stable, and comparable measurements at multiple standardized anatomical planes and establish a reproducible multiplanar method for observing CSF hydrodynamics across the craniospinal axis.

### Ventral-dorsal dual-region flow

Previous investigations of spinal CSF hydrodynamics have often treated the SAS as a global flow space. PC-MRI quantitative analysis commonly delineates an ROI encompassing the entire SAS on a single axial plane and calculates parameters such as mean and peak velocity; anatomical schematics and some computational models likewise simplify the spinal SAS as an annular compartment around the spinal cord [6, 21]. This approach facilitates measurement and comparison of global parameters but assumes that all regions within an axial plane can be regarded as one hydrodynamic unit. Such global treatment may not be applicable to the spinal SAS. Previous studies show that CSF motion is driven jointly by cardiac and respiratory cycles, and its velocity, direction, and phase may vary across spinal segments [7, 15]. In disorders such as Chiari malformation, syringomyelia, and subarachnoid stenosis, local anatomical narrowing, altered compliance, and restricted pathways may also affect CSF flow [19, 20]. Nerve roots, denticulate ligaments, arachnoid trabeculae, and pial extensions are distributed around the spinal cord and may alter local hydraulic resistance and pressure transmission [22]. Thus, although the spinal SAS is anatomically continuous, its internal flow need not be homogeneous, synchronous, or directionally uniform.

Under these circumstances, analysis based on a global ROI (a single-compartment assumption) may average the directional differences between ventral and dorsal regions [6, 7]. A global ROI may be unsuitable when the two regions exhibit longitudinal oscillations with opposite directions, different phases, or different magnitudes at the same cardiac phase. Continued use of a global ROI may yield similar mean velocities, low net flow, or nonsignificant overall differences. Such findings cannot exclude regional flow differences within the spinal SAS; instead, they may reflect cancellation of ventral and dorsal signals with opposing directions or phases during averaging. This issue also warrants consideration in clinical assessment. Symptoms and imaging findings may remain unsatisfactory in some patients with Chiari malformation, syringomyelia, or arachnoid adhesions after relief of major narrowing or reconstruction of local pathways [23, 24]. The reason may not depend only on whether the global CSF pathway has reopened; it may also relate to preoperative and postoperative assessments that do not cover craniospinal CSF circulation and treat the spinal SAS as a single annular compartment. Under this assessment model, directional differences, phase relationships, and abnormalities of local pressure transmission between ventral and dorsal regions may be averaged, underestimating local hydrodynamic factors that affect CSF circulation. Preserving ventral and dorsal directional information in the axial plane, together with multiplanar craniospinal assessment, may therefore aid more accurate etiological interpretation, explanation of postoperative effects, and subsequent treatment decisions [19, 20].

The present findings support separate analysis of ventral and dorsal regions as two principal flow regions. Multiplanar phase images showed that phase signals within the spinal SAS were not randomly distributed but formed relatively stable ventral and dorsal regions. Participant-level paired analysis showed similar mean and absolute peak velocities, indicating that the ventral and dorsal regions are not simply high- and low-flow areas. Directional peak velocity differed, suggesting different directional behavior over the cardiac cycle. Sequential cardiac-phase images further showed antiphase changes in phase signals, supporting a phase difference or directional difference between the two regions. On the basis of these findings, spinal CSF flow may comprise two relatively independent yet coordinated longitudinal oscillatory regions, ventral and dorsal. Relative independence refers to different distributions of phase signals and directional velocity characteristics within the axial plane; coordination indicates that the two regions remain within the same continuous SAS and are jointly influenced by the cardiac cycle, craniospinal pressure gradients, and changes in local compliance. Thus, ventral-dorsal dual regions should not be interpreted simply as two closed anatomical chambers; they may instead represent dynamic subdivisions within a continuous CSF space arising from local anatomical boundaries, pressure transmission, and hydraulic resistance.

A lateral fibrous-neural complex may provide an important anatomical basis for ventral-dorsal partitioning. High-resolution T2-weighted imaging (Figure 7) showed denticulate ligaments, arachnoid trabeculae, nerve roots, pial extensions, and accompanying vascular branches along the lateral aspect of the spinal cord, in the transition zones between the ventral and dorsal flow regions [22]. These structures may limit disordered mixing of ventral and dorsal CSF in the axial plane and may affect local pressure transmission through spaces around nerve roots, vascular branches, and arachnoid trabeculae [21].

**Figure 6.**
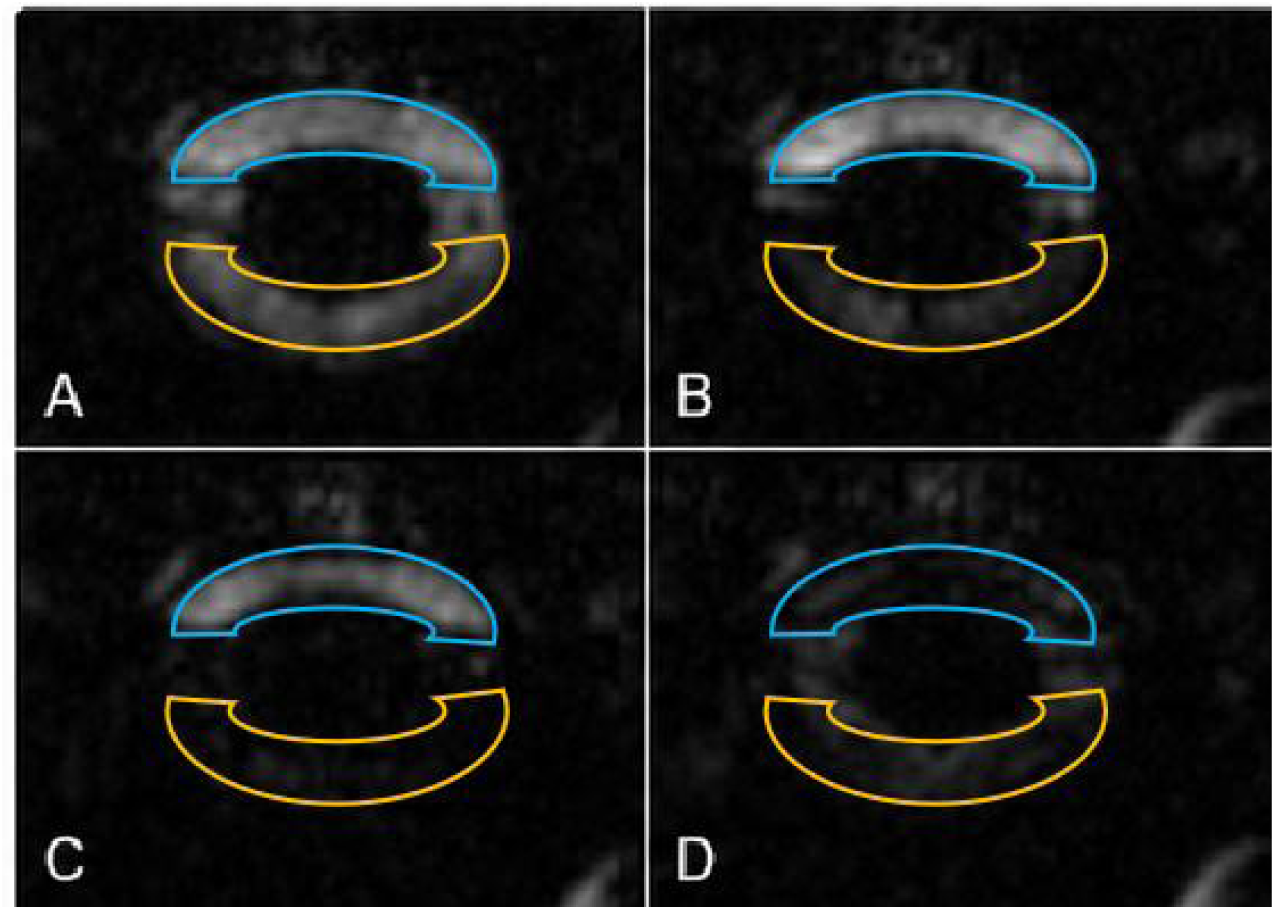
Representative sequential cardiac-phase PC-MRI images. The images show changes in phase signals within the spinal SAS over the cardiac cycle. From phase A to B, phase signals gradually increased in the ventral region and relatively decreased in the dorsal region. The distribution then shifted, with lower ventral and higher dorsal signals from phases C to D. This continuous dynamic process suggests opposite trends in the phase signals of the ventral and dorsal regions across the cardiac cycle.

**Figure 7.**
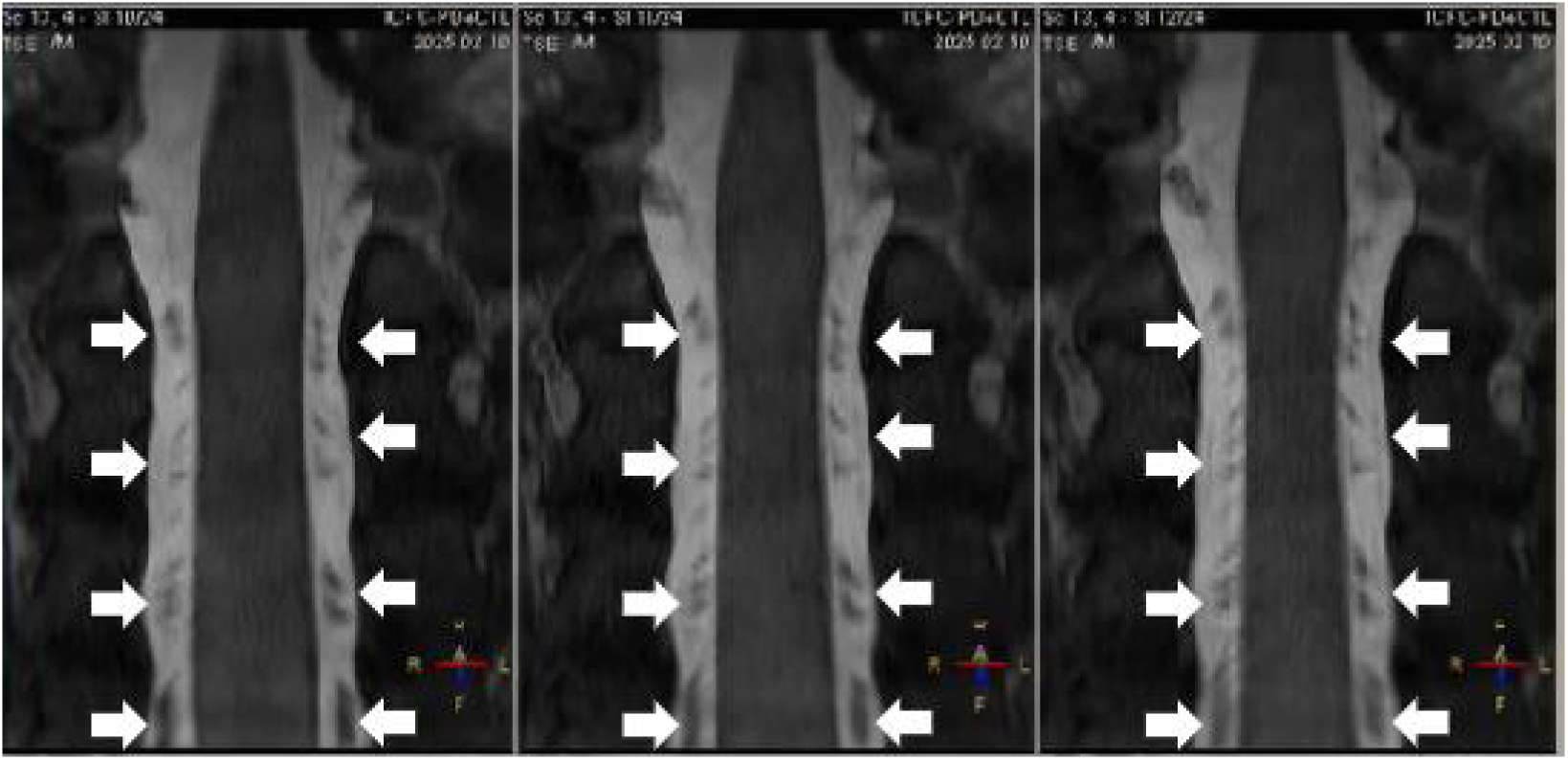
Coronal T2-weighted image of spinal anatomical structures in the fibro-neuro-radicular complex. This image illustrates the anatomical basis proposed for the fluid separation described above. White arrows indicate band-like hypointense structures extending longitudinally within the SAS. These structures comprise a fibro-neuro-radicular complex formed by spinal nerve rootlets and denticulate ligaments. This complex constitutes a natural physical barrier that partitions the continuous fluid space into ventral and dorsal flow channels.

### Potential metabolic-exchange and clinical translational implications

The value of the multiplanar craniospinal CSF hydrodynamic observation method established in this study is not limited to describing velocity changes at different planes; it also offers a perspective on the spatial basis of fluid exchange, pressure transmission, and solute transport. Recent research on meningeal lymphatic vessels, perivascular spaces, perineural pathways, and solute clearance suggests that CSF may contribute to central nervous system homeostasis through multiple interrelated systems [16-18]. The ventral-dorsal dual-region flow pattern observed here suggests that the spinal SAS is not a completely homogeneous space but contains regional partitioning. Directional differences between ventral and dorsal regions may provide anatomical-functional clues for further study of perispinal fluid exchange and solute transport.

In this context, lateral spinal nerve roots and their surrounding fibrous structures warrant further attention. If future studies confirm the presence of relatively independent yet coordinated ventral and dorsal longitudinal oscillatory regions in the spinal SAS, lateral nerve roots and perivascular regions around accompanying vessels may be candidate sites for investigating perispinal solute exchange and pressure transmission [18, 25]. However, this study did not directly measure interregional fluid exchange, solute transport, or pressure gradients. These interpretations therefore remain research hypotheses based on imaging findings and anatomical structures and require further investigation using tracer imaging, real-time respiratory-synchronized imaging, 4D Flow MRI, or computational fluid dynamics [18, 25].

This observational framework may also provide more refined imaging phenotypes for mechanistic research on disorders related to CSF hydrodynamics. Chiari malformation, syringomyelia, hydrocephalus, arachnoid adhesions, post-traumatic spinal adhesions, and postoperative scars may all alter SAS morphology, local compliance, and spaces around nerve roots, thereby affecting CSF flow characteristics in different regions [19, 20, 26]. If only global-ROI velocity or flow is evaluated, local signals with opposing directions may be partially canceled and regional changes may be missed [7, 10]. Ventral-dorsal partitioning analysis could complement conventional global-ROI measurements by comparing whether regional flow patterns change in disease states and by further exploring their associations with clinical symptoms, structural imaging changes, and disease progression.

In treatment research, this method could be used to observe changes in regional CSF flow characteristics before and after decompression surgery, shunt treatment, and other interventions, providing supplementary imaging information for efficacy assessment and mechanistic research. For example, if global velocity changes little after an intervention while directional parameters in ventral or dorsal regions change, this may indicate adjustment of the local flow environment or pressure-transmission pattern. Whether such changes are associated with symptom improvement or long-term outcomes requires validation in longitudinal follow-up studies. Ventral-dorsal partitioning may also affect evaluation of intrathecal drug delivery and local infusion therapy. Current estimates of intrathecal drug distribution usually consider injection location, CSF volume, global velocity, and drug physicochemical properties [27]. Given regional flow patterns with different directions in the spinal SAS, the distribution of drugs or tracers around the spinal cord may also be influenced by local flow direction, cardiac-cycle phase, segmental location, and structures around nerve roots [28]. Ventral-dorsal partitioning parameters may therefore serve as candidate observational measures in future studies of intrathecal drug delivery, CSF replacement, local perfusion, and neuromodulation-related treatments to explore regional distribution of drugs or tracers.

## Conclusions

Using electrocardiography-gated two-dimensional Cine PC-MRI and a uniform Q-Flow post-processing workflow, this study performed multiplanar, quantitative, and dynamic observations of CSF flow in healthy adults at six standardized anatomical levels from the cerebral aqueduct outlet/fourth-ventricle inlet to the lumbar cistern. The results showed that: (1) CSF flow within the spinal SAS had clear regional ventral-dorsal characteristics. Mean velocity and absolute peak velocity were similar in the two regions, whereas directional peak velocity differed (1.50 ± 2.98 cm/s vs. -0.38 ± 3.23 cm/s, P = 0.036); sequential cardiac-phase images also showed changes in signals in the two regions across the cardiac cycle. (2) These findings do not support a simple view of the spinal SAS as a homogeneous, synchronous, single annular flow space. Global ROI analysis may cancel local velocity signals with opposing directions and thereby obscure regional flow characteristics.

The standardized multiplanar Cine PC-MRI observation strategy established here allows noninvasive, repeatable, and comparable quantitative analysis of CSF flow across the craniospinal axis. It provides an initial description of craniospinal CSF hydrodynamic characteristics in healthy individuals and an exploratory imaging reference for CSF hydrodynamics. This method may be further applied in Chiari malformation, syringomyelia, hydrocephalus, and arachnoid adhesions to compare regional flow patterns, phase relationships, and changes before and after intervention, providing a methodological basis for mechanistic research and efficacy evaluation.

## Data Availability

All data produced in the present study are available upon reasonable request to the authors

## List of abbreviations

CSF: cerebrospinal fluid
CNS: central nervous system
SAS: subarachnoid space
MRI: magnetic resonance imaging
PC-MRI: phase-contrast magnetic resonance imaging
Cine PC-MRI: cine phase-contrast magnetic resonance imaging
ROI: region of interest
ECG: electrocardiogram
ICC: intraclass correlation coefficient
CFD: computational fluid dynamics
TR: repetition time
TE: echo time
VENC: velocity encoding.

## Declarations

### Ethics approval and consent to participate

The study was approved by the Medical Ethics Committee of Tsinghua University Yuquan Hospital (approval no. 2024KY014). Written informed consent was obtained from all participants.

### Consent for publication

Not applicable. This article does not contain any individually identifiable data or images.

### Availability of data and materials

The de-identified data supporting the conclusions of this article are available from the corresponding author on reasonable request, subject to institutional and ethical restrictions.

### Authors’ contributions

Liying Sun performed the experimental procedures, acquired and analyzed the data, and drafted the manuscript. Le He provided expert guidance on MRI acquisition and image analysis. Zhimo Jian contributed to statistical analysis, data analysis, and interpretation of the results. Tiange Lu, Tiemin Li, and Mingyuan Yan assisted with statistical and data analyses. Suhua Miao contributed to experimental design and data analysis. Rongsong Zhou performed MRI scanning and participated in data acquisition and analysis. Yajun Yin provided expertise in mechanics and guidance on experimental design, interpretation of the experimental data, research direction, and formulation of the conclusions. Yu Ma led the overall study and was responsible for overall experimental design, study coordination, discussion and interpretation of the results, and formulation of the conclusions. Yuqi Zhang contributed to overall experimental design and provided guidance on data analysis. All authors read and approved the final manuscript.

## Acknowledgements

Not applicable.

## References

1. Hladky SB, Barrand MA. Regulation of brain fluid volumes and pressures: basic principles, intracranial hypertension, ventriculomegaly and hydrocephalus. Fluids Barriers CNS. 2024;21:57. doi:10.1186/s12987-024-00532-w.

2. Xiang J, Hua Y, Xi G, Keep RF. Mechanisms of cerebrospinal fluid and brain interstitial fluid production. Neurobiol Dis. 2023;183:106159. doi:10.1016/j.nbd.2023.106159.

3. Al Masri O, Mouchtouris N, Ashraf O, et al. Mapping cerebrospinal fluid dynamics in central nervous system disorders: a scoping review. Neurosurg Rev. 2024;47:136. doi:10.1007/s10143-024-02291-6.

4. Brinker T, Stopa E, Morrison J, Klinge P. A new look at cerebrospinal fluid circulation. Fluids Barriers CNS. 2014;11:10. doi:10.1186/2045-8118-11-10.

5. Atchley TJ, Vukic B, Vukic M, Walters BC. Review of cerebrospinal fluid physiology and dynamics: a call for medical education reform. Neurosurgery. 2022;91:1–7. doi:10.1227/neu.0000000000002000.

6. Korbecki A, Zimny A, Podgórski P, Sąsiadek M, Bladowska J. Imaging of cerebrospinal fluid flow: fundamentals, techniques, and clinical applications of phase-contrast magnetic resonance imaging. Pol J Radiol. 2019;84:e240–e250. doi:10.5114/pjr.2019.86881.

7. Eide PK, Valnes LM, Lindstrøm EK, Mardal KA, Ringstad G. Direction and magnitude of cerebrospinal fluid flow vary substantially across central nervous system diseases. Fluids Barriers CNS. 2021;18:16. doi:10.1186/s12987-021-00251-6.

8. Chen Y, Hong H, Nazeri A, Markus HS, Luo X. Cerebrospinal fluid-based spatial statistics: towards quantitative analysis of cerebrospinal fluid pseudodiffusivity. Fluids Barriers CNS. 2024;21:59. doi:10.1186/s12987-024-00559-z.

9. Vikner T, Johnson KM, Cadman RV, Betthauser TJ, Wilson RE, Chin N, et al. CSF dynamics throughout the ventricular system using 4D flow MRI: associations to arterial pulsatility, ventricular volumes, and age. Fluids Barriers CNS. 2024;21:68. doi:10.1186/s12987-024-00570-4.

10. Williams G, Thyagaraj S, Fu A, Loth F, Oshinski JN, Haughton V, et al. In vitro evaluation of cerebrospinal fluid velocity measurement in type I Chiari malformation: repeatability, reproducibility, and agreement using 2D phase contrast and 4D flow MRI. Fluids Barriers CNS. 2021;18:12. doi:10.1186/s12987-021-00246-3.

11. Yamada S, Otani T, Ii S, Ito H, Iseki C, Tanikawa M, et al. Modeling cerebrospinal fluid dynamics across the entire intracranial space through integration of four-dimensional flow and intravoxel incoherent motion magnetic resonance imaging. Fluids Barriers CNS. 2024;21:47. doi:10.1186/s12987-024-00552-6.

12. Wright AM, Wu YC, Feng L, Wen Q. Diffusion magnetic resonance imaging of cerebrospinal fluid dynamics: current techniques and future advancements. NMR Biomed. 2024;37:e5162. doi:10.1002/nbm.5162.

13. Yamada S, Hiratsuka S, Otani T, et al. Usefulness of intravoxel incoherent motion MRI for visualizing slow cerebrospinal fluid motion. Fluids Barriers CNS. 2023;20:16. doi:10.1186/s12987-023-00415-6.

14. Feinberg DA, Mark AS. Human brain motion and cerebrospinal fluid circulation demonstrated with MR velocity imaging. Radiology. 1987;163:793–799. doi:10.1148/radiology.163.3.3575734.

15. Fultz NE, Bonmassar G, Setsompop K, Stickgold RA, Rosen BR, Polimeni JR, et al. Coupled electrophysiological, hemodynamic, and cerebrospinal fluid oscillations in human sleep. Science. 2019;366:628–631. doi:10.1126/science.aax5440.

16. Iliff JJ, Wang M, Liao Y, Plogg BA, Peng W, Gundersen GA, et al. A paravascular pathway facilitates CSF flow through the brain parenchyma and the clearance of interstitial solutes, including amyloid β. Sci Transl Med. 2012;4:147ra111. doi:10.1126/scitranslmed.3003748.

17. Louveau A, Smirnov I, Keyes TJ, Eccles JD, Rouhani SJ, Peske JD, et al. Structural and functional features of central nervous system lymphatic vessels. Nature. 2015;523:337–341. doi:10.1038/nature14432.

18. Ligocki AP, Vinson AV, Yachnis AT, Dunn WA Jr, Smith DE, Scott EA, et al. Cerebrospinal fluid flow extends to peripheral nerves further unifying the nervous system. Sci Adv. 2024;10:eadn3259. doi:10.1126/sciadv.adn3259.

19. Bhadelia RA, Chang YM, Oshinski JN, Loth F. Cerebrospinal fluid flow and brain motion in Chiari I malformation: past, present, and future. J Magn Reson Imaging. 2023;58:360–378. doi:10.1002/jmri.28717.

20. Heiss JD. Cerebrospinal fluid hydrodynamics in Chiari I malformation and syringomyelia: modeling pathophysiology. Neurosurg Clin N Am. 2023;34:81–90. doi:10.1016/j.nec.2022.08.007.

21. Putluru DR, Buganza Tepole A, Gomez H. Mixed-dimensional fluid-structure interaction simulations reveal key mechanisms of cerebrospinal fluid dynamics in the spinal canal. Fluids Barriers CNS. 2025;22:81. doi:10.1186/s12987-025-00691-4.

22. Heidari Pahlavian S, Yiallourou T, Tubbs RS, Bunck AC, Loth F, Goodin M, et al. The impact of spinal cord nerve roots and denticulate ligaments on cerebrospinal fluid dynamics in the cervical spine. PLoS One. 2014;9:e91888. doi:10.1371/journal.pone.0091888.

23. Oldfield EH, Muraszko K, Shawker TH, Patronas NJ. Pathophysiology of syringomyelia associated with Chiari I malformation of the cerebellar tonsils: implications for diagnosis and treatment. J Neurosurg. 1994;80:3–15. doi:10.3171/jns.1994.80.1.0003.

24. Bhadelia RA, Bogdan AR, Wolpert SM, Lev S, Appignani BA, Heilman CB. Cerebrospinal fluid flow waveforms: analysis in patients with Chiari I malformation by means of gated phase-contrast MR imaging velocity measurements. Radiology. 1995;196:195–202. doi:10.1148/radiology.196.1.7784567.

25. Boster KAS, Sun J, Shang JK, Kelley DH, Thomas JH. Hydraulic resistance of three-dimensional pial perivascular spaces in the brain. Fluids Barriers CNS. 2024;21:7. doi:10.1186/s12987-023-00505-5.

26. Williams MA, Nagel SJ, Golomb J, Jensen H, Dasher NA, Holubkov R, et al. Safety and effectiveness of the assessment and treatment of idiopathic normal pressure hydrocephalus in the Adult Hydrocephalus Clinical Research Network. J Neurosurg. 2022;137:1289–1301. doi:10.3171/2022.1.JNS212782.

27. Haga PT, Pizzichelli G, Mortensen M, Kuchta M, Heidari Pahlavian S, Sinibaldi E, et al. A numerical investigation of intrathecal isobaric drug dispersion within the cervical subarachnoid space. PLoS One. 2017;12:e0173680. doi:10.1371/journal.pone.0173680.

28. Parras-Martos FJ, Sánchez AL, Martínez-Bazán C, Coenen W, Gutiérrez-Montes C. On reduced-order modeling of drug dispersion in the spinal canal. Fluids Barriers CNS. 2025;22:66. doi:10.1186/s12987-025-00657-6.

29. Pripp AH, Ringstad G, Valnes LM, Eide PK. A method for measuring first glymphatic influx of a cerebrospinal fluid tracer in the human brain. Front Neurosci. 2025;19:1703748. doi:10.3389/fnins.2025.1703748.

30. Boyd ED, Kaur J, Ding G, Chopp M, Jiang Q. Clinical magnetic resonance imaging evaluation of glymphatic function. NMR Biomed. 2024;37:e5132. doi:10.1002/nbm.5132.

31. Ringstad G, Eide PK. Glymphatic MRI in idiopathic normal pressure hydrocephalus. Cell Mol Life Sci. 2024;81:25. doi:10.1007/s00018-023-05044-w.

32. Jiang-Xie LF, Drieu A, Bhasin K, Quintero D, Smirnov I, Kipnis J. Neuronal dynamics direct cerebrospinal fluid perfusion and brain clearance. Nature. 2024;627:157–164. doi:10.1038/s41586-024-07108-6.

33. Rowsthorn E, Pham W, Nazem-Zadeh MR, Law M, Pase MP, Harding IH. Imaging the neurovascular unit in health and neurodegeneration: a scoping review of interdependencies between MRI measures. Fluids Barriers CNS. 2023;20:97. doi:10.1186/s12987-023-00499-0.

34. Miao A, Luo T, Hsieh B, Edge CJ, Gridley M, Wong RTC, et al. Brain clearance is reduced during sleep and anesthesia. Nat Neurosci. 2024;27:1046–1050. doi:10.1038/s41593-024-01638-y.

35. Keep RF, Jones HC, Drewes LR. Advances in brain barriers and brain fluids research in 2021: great progress in a time of adversity. Fluids Barriers CNS. 2022;19:48. doi:10.1186/s12987-022-00343-x.

